# Deep Learning Using High-Resolution Images of Forearm Predicts Fracture

**DOI:** 10.1101/2023.04.05.23288167

**Authors:** Roland Chapurlat, Serge Ferrari, Xiaoxu Li, Yu Peng, Min Xu, Min Bui, Elisabeth Sornay-Rendu, Eric lespessailles, Emmanuel Biver, Ego Seeman

## Abstract

**Importance:** Fragility fractures are a public health problem. Over 70% of women having fractures have osteopenia or normal BMD, but they remain unidentified and untreated because the definition of ‘osteoporosis’, a bone mineral density (BMD) T-Score ≤ -2.5SD, is often used to signal bone fragility.

**Objective:** As deep learning facilitates investigation of bone’s multi-level hierarchical structure and soft tissue, we tested whether this approach might better identify women at risk of fracture before fracture.

**Design:** We pooled data from three French and Swiss prospective population-based cohorts (OFELY, QUALYOR, GERICO) that collected clinical risk factors for fracture, areal BMD and distal radius measurements with high resolution peripheral quantitative tomography (HRpQCT). Using only three-dimensional images of the distal radius, ulna and soft tissue acquired by HRpQCT, an algorithm, a Structural Fragility Score-Artificial Intelligence (SFS-AI), was trained to distinguish 277 women having fractures from 1401 remaining fracture-free during 5 years and then was tested in a validation cohort of 422 women.

**Setting:** European postmenopausal women

**Participants:** We have studied postmenopausal women considered as representative of the general population, who were followed for a median 9.4 years in OFELY, 5.4 years in QUALYOR and 5.7 years in GERICO.

**Main outcome and measure:** All types of incident fragility fractures

**Results:** We used data from 2666 postmenopausal women, with age range of 42-94. In women ≥ 65 years having ‘All Fragility Fractures’ or ‘Major Fragility Fractures’, SFS-AI generated an AUC of 66-70%, sensitivities of 60-68% and specificity of 71%. Sensitivities were greater than achieved by the fracture risk assessment (FRAX) with BMD or BMD (6.7-26.7%) with lower specificities than these diagnostics (∼95%).

**Conclusion and relevance:** The SFS-AI is a holistic surrogate of fracture risk that pre-emptively identifies most women needing prompt treatment to avert a first fracture.

**Key Points:** *Question:* Can a deep learning model (DL)° based on high resolution images of the distal forearm predict fragility fractures?

*Findings:* In the setting of 3 pooled population-based cohorts, the DL model predicted fractures substantially better than areal bone mineral density and FRAX, especially in women ≥65 years.

*Meaning:* Our DL model may become an easy to use way to identify postmenopausal women at risk for fracture to improve fracture prevention.

## Introduction

Fragility fractures are a public health problem because fractures impose high morbidity, mortality and cost to the community.^1^ To identify women with fragile bones before fracture, a W.H.O group designated women as having ‘osteoporosis’ if femoral neck bone mineral density (BMD) T-Score was ≤ -2.5 standardized deviations (SD) below the premenopausal mean.^2^ Epidemiological studies confirmed that fracture risk increases as BMD decreases, but the frequency distribution around the age-related decline in mean BMD remains normal.

Because of this normal frequency distribution, most postmenopausal women in the community have osteopenia (T-score -2.5 to -1.0 SD) or normal BMD (T-score > -1.0 SD). These women form the source of 75% of all fragility in the community, only 25% arise among the smaller subset of women in the community with osteoporosis as defined by BMD.^3-9^ The women with osteopenia or normal BMD having fragility fractures remain unidentified and untreated using the definition of osteoporosis, a BMD T-Score ≤ - 2.5 SD, to signal bone fragility. Treatment is not offered, even in the presence of a prevalent or incident fracture, because the absence of osteoporosis is incorrectly interpreted as being evidence of absence of bone fragility.^10^

Osteoporosis and bone fragility are used interchangeably even though they are not synonymous terms.^11-13^ Absence of osteoporosis does not exclude bone fragility. Bone fragility is not binary, present in women with osteoporosis (T-Score ≤ -2.5 SD) and absent in women without osteoporosis (T-Score > -2.5 SD). Even when bone loss only reduces BMD into the low normal or osteopenia range, the bone is unlikely to be ‘normal’. Bone mass is reduced relative to premenopausal women and many qualities of bone responsible for its strength may be compromised.^14-18^

For example, advancing age deteriorates the composition of the mineralized matrix.^19,20^ Bone loss disrupts the spatial configuration of bone’s three-dimensional architecture.^15,21^ These changes produce a non-linear increase in bone fragility, disproportionate to both the bone loss causing the deterioration and the reduction in BMD.^20,22^ Resistance to bending is a 7^th^ power function of bone’s cortical porosity and a 3^rd^ power function of its trabecular density.^22^

Consequently, even modest disruption of the spatial configuration of bone at nano-, and micro-levels of resolution compromise bone strength independent of BMD. In addition, soft tissue changes like loss of muscle mass (sarcopenia) impair mobility and balance predisposing to falls, fractures and mortality.^23^ Thus, reducing the population burden of fractures requires a diagnostic that complements BMD by identifying women at risk of fracture due to bone fragility caused by compromised bone morphology not captured by BMD ≤ - 2.5 SD, by an increased risk of falls due to deteriorated soft tissues such as muscle mass, or both. Non-invasive evaluation of bone microarchitecture improves fracture prediction compared with FRAX plus BMD or BMD alone.^8,9^

A promising area of innovation in the promotion of human health is the use of Artificial Intelligence (AI). Application of Deep Learning to medical imaging^24^ facilitates the investigation of bone’s multilayered qualities and has been reported to identify patients with prevalent fractures or osteoporosis in cross sectional studies.^25-27^ However, no prospective studies have applied deep learning using only the high resolution 3-dimensional images of bone and soft tissue to determine whether an algorithm, a Structural Fragility Score derived by Artificial Intelligence (SFS-AI), might capture deteriorated bone qualities and soft tissue. If so, this holistic surrogate of fracture risk is likely to serve as a diagnostic that pre-emptively identifies women at risk of a first or subsequent fracture needing prompt treatment and would do so better than the fracture risk assessment (FRAX) score with BMD or BMD alone.

## Methods

### Participants

We studied (i) 568 postmenopausal women, median age 68.2 years, range 42-94 of Os des FEmmes de LYon, OFELY, France followed for a median [interquartile range] of 9.4 [1.0] years,^9,28,29^ (ii) 1427 women of the Qualité Osseuse Lyon Orléans, QUALYOR cohort (1042 recruited in Lyon, 497 in Orléans), median age 65.9 (range 50-87) years followed for 5 years^30^ and (iii) 671 women of the Geneva Retirees Cohort, GERICO, in Switzerland median age 65 (range 63-68) years followed 5.7 years (range 2-8).^31^ The studies were approved by the institutional review boards. Participants provided informed consent. Fractures (excluding head, toes and fingers) were confirmed using radiographs.

### Bone microarchitecture, bone densitometry, FRAX with BMD

HRpQCT (voxel size of 82 μm^3^) was used to scan the non-dominant forearm (Scanco Medical AG, Switzerland).^32^ Radiation exposure was ∼3 microsievert. Quality control was monitored by daily scans of hydroxyapatite rods (QRM, Moehrendorf, Germany). Femoral neck BMD was quantified using Hologic DXA scanners in the French cohorts and Hologic QDR Discovery in the Swiss cohort. T-scores were calculated using NHANES III. FRAX with femoral neck BMD provides a 10-year risk for Major Fragility Fractures (proximal humerus, wrist, distal forearm, clinical spine, or hip).^33^

### Deep Learning network

To avoid bias towards any one of the three cohorts, we combined the three cohorts and then we randomly divided the combined data set into a training and testing data set. No training data was used as testing data. **Figure S2** shows participants were randomly allocated to five groups, four used for training (n=1678), the fifth used for testing (n = 422). Scores were calculated for each testing group with the median forming the SFS-AI (see Supplement **Figure S2**). Deep learning was applied to images of the distal radius and ulna and the surrounding soft tissue acquired using HR-pQCT (see Supplement).^34-37^ Training the algorithm to identify women sustaining fractures faced two challenges: (i) extraction of features within the three-dimensional image captured by a matrix of 110*1560*1560 voxels conferring fracture risk and (ii) limited data for training predisposing to model over-fitting. We used the DenseNet121 as the feature extraction network (**Figure S1**). Features conferring fracture risk were learnt collectively by densely connected layers in the neural network. The input to the feature extraction network was the 110 slices acquired by the HR-pQCT at the distal radius (including the ulna and surrounding soft tissues). The output from the feature extraction network is a feature vector of 256 numbers.

To achieve robust feature extraction, a multi-task learning strategy was used to overcome model overfitting. To provide pictorial representation of the fracture risk prediction, extracted features were displayed as a heat map overlaid upon a 2D projection of the images. Red reflects greater relevance of the region’s bone or soft tissue to fracture prediction.

### Statistical Analyses

Analyses were conducted using data in women of any age and those 65 years and over. Follow-up was to fracture or freedom from fracture for five years since HR-pQCT scanning. SFS-AI, FRAX with BMD and BMD were not normally distributed and so are presented as median and interquartile range (IQR). Values are adjusted for age and cohort because of cohort differences in age and follow-up duration. (**Tables S1** and **S2**.) Comparison of the diagnostics in women having fractures and those remaining fracture-free was carried out using analysis of covariance adjusted for age and cohort and estimated by robust regression. (**Table 1**.)

**Table 1.**
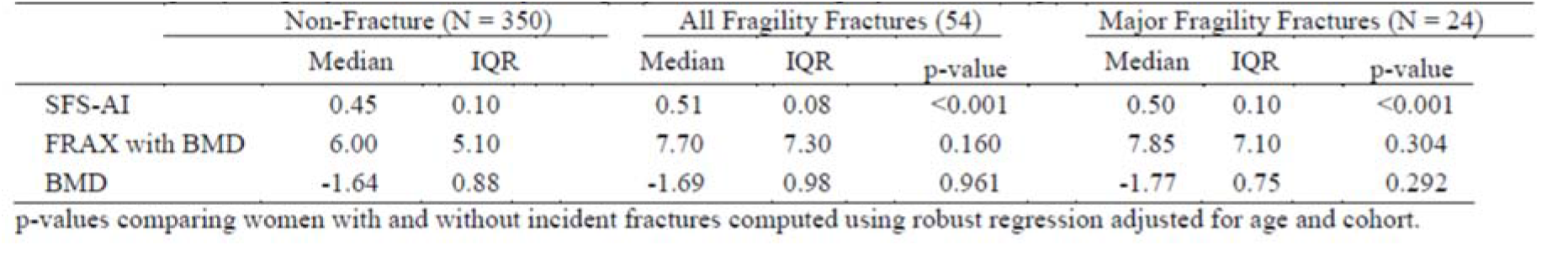
Median and interquartile ranges (IQR) for Structural Fragility Score-Artificial Intelligence (SFS-AI). Fracture Risk Assessment score (FRAX) with bone mineral density (BMD) and femoral neck BMD in women remaining fracture-free and women having Any Fragility Fractures or Major Fragility Fractures during 5 years follow-up.

|  | Non-Fracture (N = 350) |  | All Fragility Fractures (54) |  |  | Major Fragility Fractures (N = 24) |  |  |
| --- | --- | --- | --- | --- | --- | --- | --- | --- |
|  | Median | IQR | Median | IQR | p-value | Median | IQR | p-value |
| SFS-AI | 0.45 | 0.10 | 0.51 | 0.08 | <0.001 | 0.50 | 0.10 | <0.001 |
| FRAX with BMD | 6.00 | 5.10 | 7.70 | 7.30 | 0.160 | 7.85 | 7.10 | 0.304 |
| BMD | -1.64 | 0.88 | -1.69 | 0.98 | 0.961 | -1.77 | 0.75 | 0.292 |

p-values comparing women with and without incident fractures computed using robust regression adjusted for age and cohort.

The performance of SFS-AI as a continuous trait was assessed using the area under the curve (AUC) and was estimated using a parametric probit model^38^ and logistic regression to derive Odds Ratios (ORs) for fracture. Both analyses are presented unadjusted and adjusted for age and cohort effect. The sensitivity and specificity of SFS-AI as a binary trait used a threshold of 0.5. (Addressed in **Table 2**.)

**Table 2.** Performance of the Structural Fragility Score-Artificial Intelligence (SFS-AI) using data in women of any age and women aged 65 years and over

|  |  | All women of any age |  |  |  | Women aged 65 and older |  |  |  |
| --- | --- | --- | --- | --- | --- | --- | --- | --- | --- |
|  |  | Any Fragility Fracture<br>(N = 422) |  | Major Fragility Fracture<br>(N = 383) |  | Any Fragility Fracture<br>(N = 236) |  | Major Fragility Fracture<br>(N = 208) |  |
|  |  | Est | 95% CI | Est | 95% CI | Est | 95% CI | Est | 95% CI |
| AUC | Unadjusted | 0.78 | (0.73, 0.84) | 0.76 | (0.67, 0.85) | 0.78 | (0.71, 0.85) | 0.76 | (0.63, 0.88) |
|  | Adjusted | 0.74 | (0.69, 0.79) | 0.73 | (0.65, 0.83) | 0.73 | (0.67, 0.80) | 0.73 | (0.61, 0.84) |
| OR | Unadjusted | 3.44 | (2.47, 4.78) | 2.96 | (1.85, 4.76) | 3.21 | (2.13, 4.83) | 2.82 | (1.55, 5.14) |
|  | Adjusted | 2.67 | (2.03, 3.51) | 2.53 | (1.68, 3.82) | 2.64 | (1.86, 3.75) | 2.54 | (1.49, 4.34) |
| Sensitivity |  | 65.7% | (53.4%, 76.7%) | 58.1% | (39.1%, 75.5%) | 74.0% | (59.7%, 85.4%) | 68.2% | (45.1%, 86.1%) |
| Specificity |  | 77.3% | (72.5%, 81.5%) | 77.3% | (72.5%, 81.5%) | 71.0% | (63.9%, 77.4%) | 71.0% | (63.9%, 77.4%) |

N= Sample size; Est = estimate; CI = confidence interval; Area under the curve (AUC) and odds ratios (OR) estimated with SFS-AI considered as continuous measurement and unadjusted and adjusted for age and cohort effect; Sensitivity and Specificity computed using clinical cut-off point of $\geq 0.5$ .

We also assessed the performance of FRAX with BMD and BMD as continuous traits using ROC analysis and computed sensitivity and specificity using thresholds of 20% for FRAX with BMD and – 2.5 SD for BMD denoting high fracture risk. Logistic regression was then used to assess any association of these diagnostics with fracture, separately and combined, for women of any age and women aged 65 years and over. (Addressed in **Figure 1 and Table S3**.)

**Figure 1.**
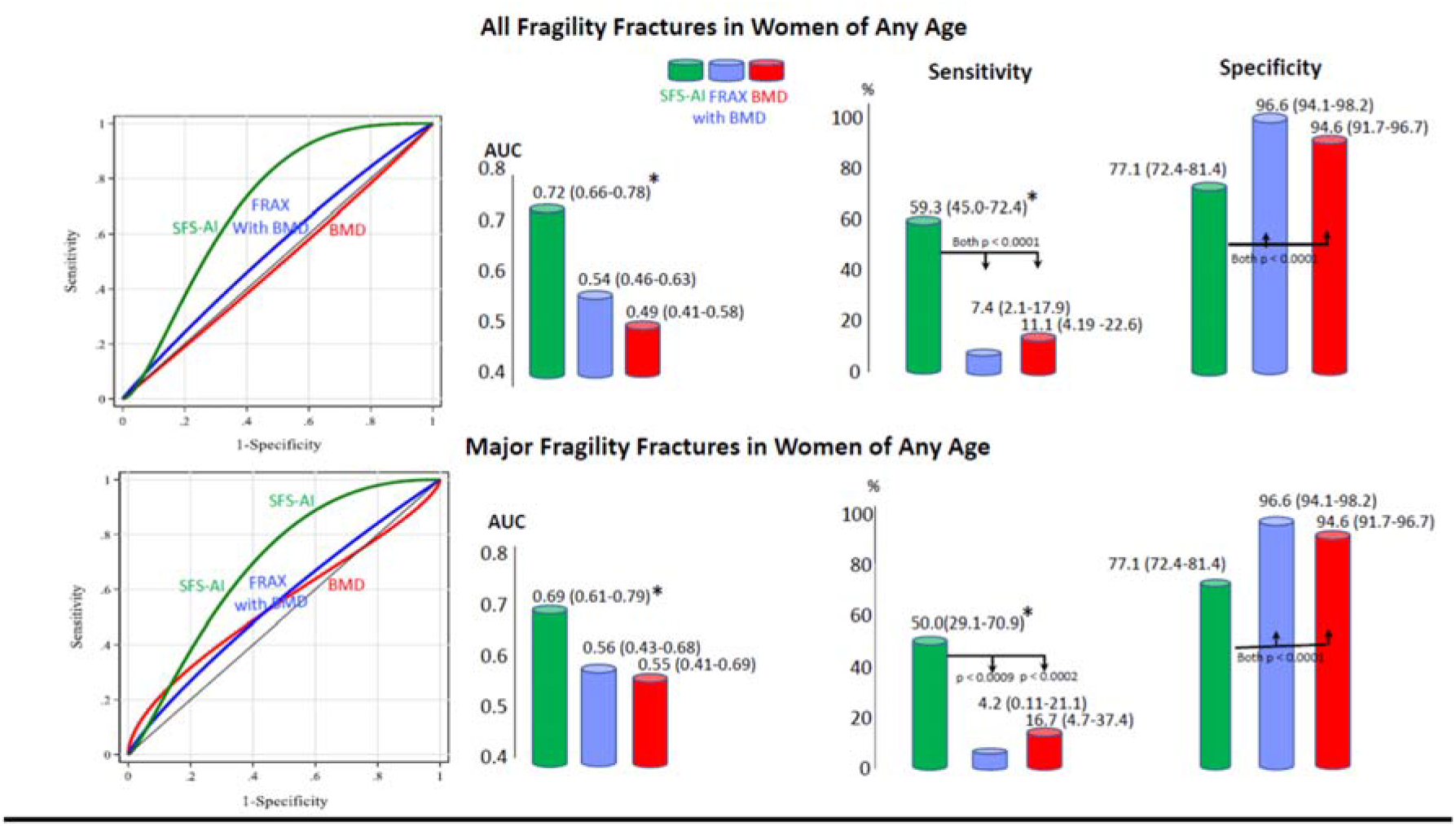
**Left two panels:** Receiver Operator Characteristic (ROC) curves for Structural Fragility Score Artificial Intelligence (SFS-AI), Fracture Risk Assessment Score (FRAX) with bone mineral density (BMD) and BMD as a continuous trait predicting for ‘All Fragility Fractures’ and ‘Majority Fragility Fractures’ for women of any age. Area under the Curves (AUCs) with 95% Confidence Intervals (CI) were significant (*p < 0.05) for SFS-AI only. **Right two panels:** Sensitivity and specificity of SFS-AI, FRAX with BMD and BMD as categorical traits.

Linear regression was used to assess the association of SFS-AI with age, separately for women with fractures and women remaining fracture-free. (Addressed in **Figure 2**.) Age, cortical porosity, trabecular density, FRAX with BMD and BMD were used in linear regression to compute an overall R-squared and to determine the proportion of variance in SFS-AI explained by these independent variables. The percentage contribution of each trait to the overall R-squared was computed using the Shapley method.^39^ (Addressed in **Table S5** and **Figure 3**.)

**Figure 2.**
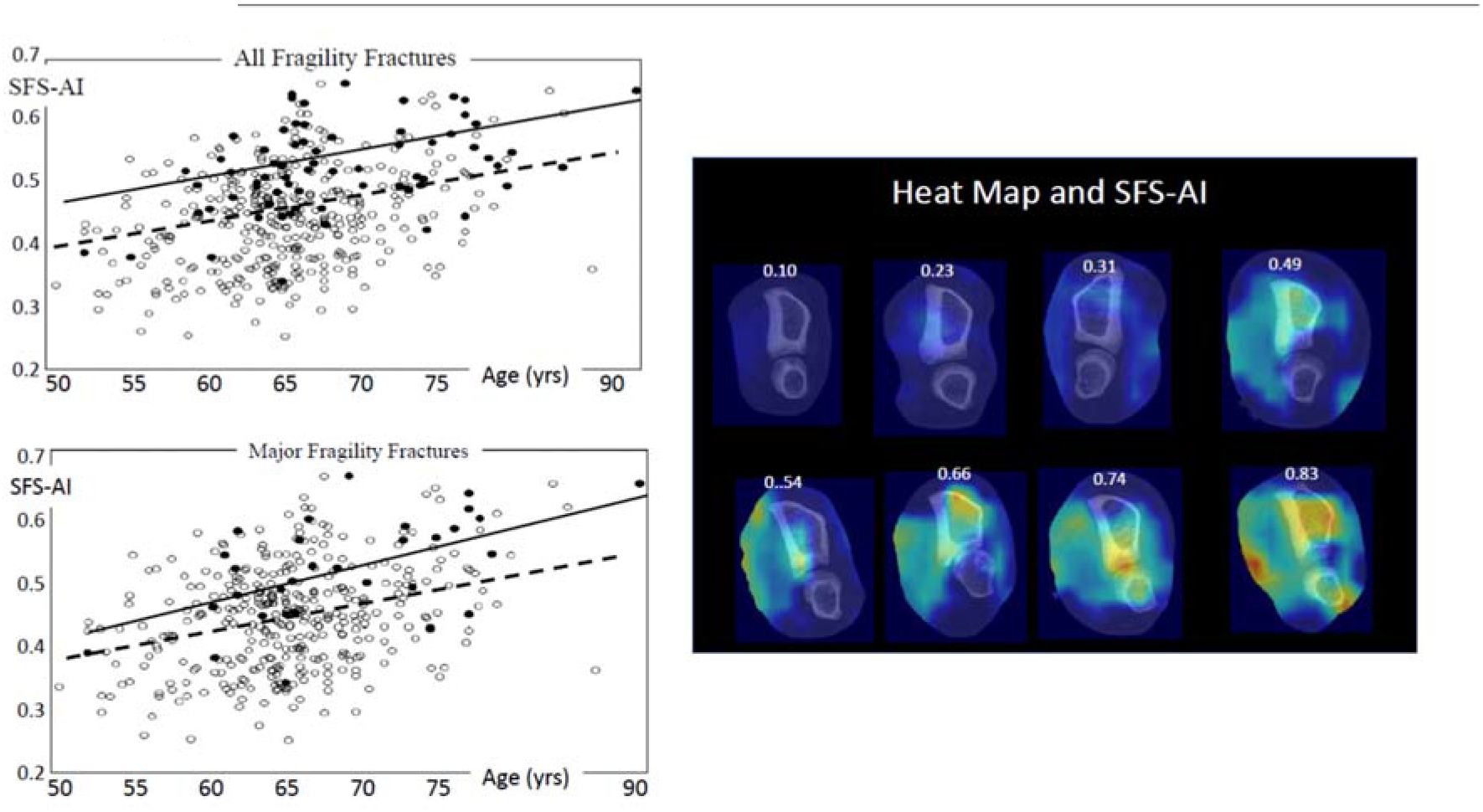
**Left panels:** Advancing age is associated with a higher Structural Fragility Score-Artificial Intelligence (SFS-AI) in women having ‘All Fragility Fractures’ or Major Fragility Fractures (closed circles) and in women remaining fracture-free (open circles). The images of the distal radius and ulna with the heat map illustrate regions commonly encountered in women having fractures.

**Figure 3.**
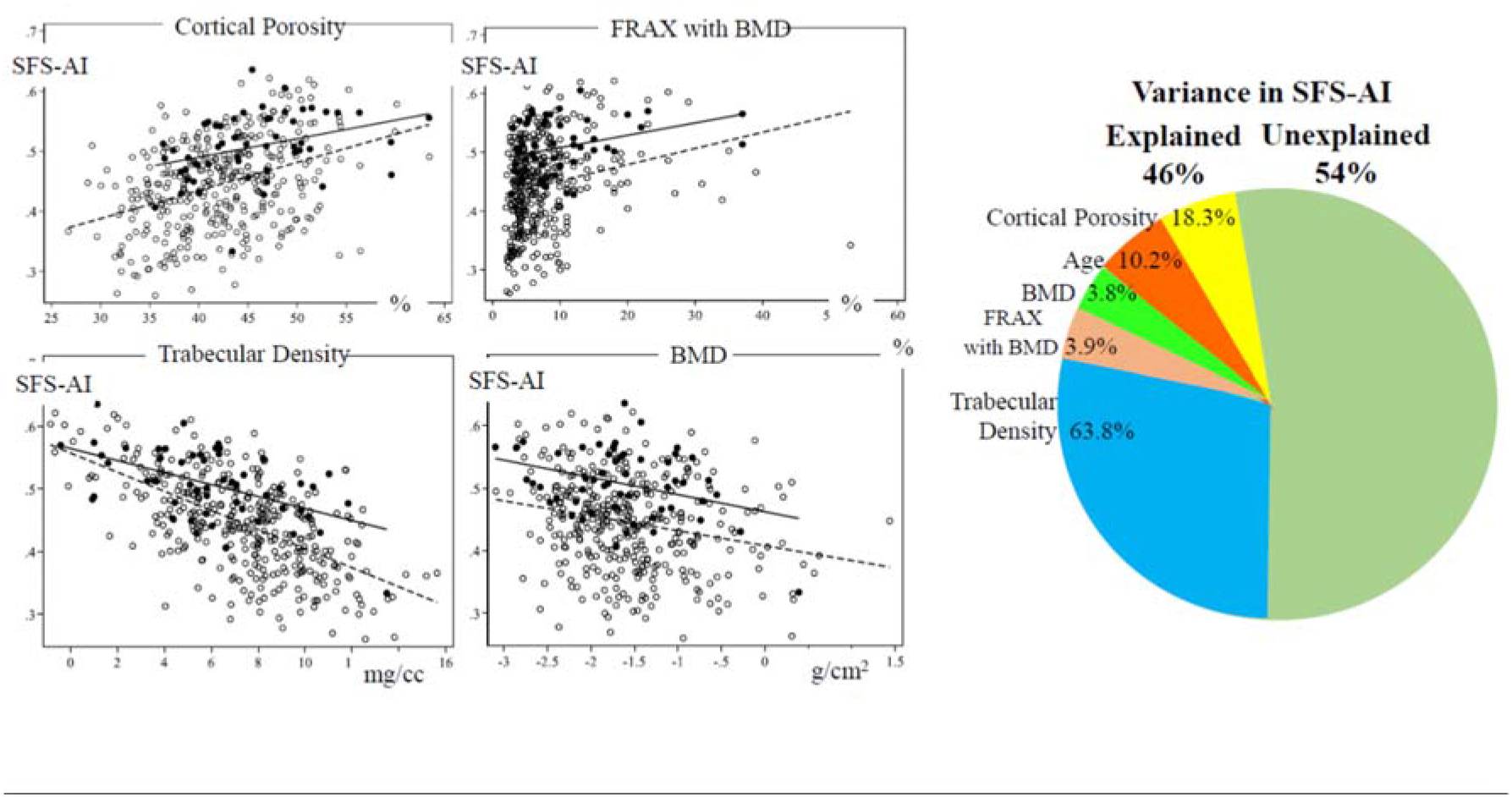
**Left panels.** The Structural Fragility Score-Artificial Intelligence (SFS-AI) was associated directly with cortical porosity, FRAX with BMD and negatively with trabecular density and BMD. **Right diagram**. Of the 47% of explained variance in the SFS-AI, most was attributed to trabecular density, cortical porosity, age and the FRAX with BMD. The contribution of BMD was not significant. The remaining 53 percent remained unexplained.

## Results

**Table 1** shows SFS-AI was higher in women having ‘All’ or ‘Major Fragility Fractures’ than women remaining fracture-free (both p < 0.001). Neither FRAX with BMD nor BMD alone differed in women having fractures versus those remaining fracture-free (p > 0.15).

### SFS-AI pre-emptively identifies women at risk of ‘All’ and ‘Major Fragility Fractures’

**Table 2** shows that in the testing cohort of 422 women of any age and the 236 women ≥ 65 years of age, the SFS-AI as a continuous trait generated AUCs of 73-74% for ‘All Fragility Fractures’ and ‘Major Fragility fractures’ with adjusted ORs ranging from 2.53 to 2.67 per standard deviation. The SFS-AI as a categorical trait (using a threshold of 0.5), had sensitivities ranging from 58.1% to 74.0% and specificities ranging from 71.0% to 77.3% (all significant, p < 0.001 for OR and AUC.

### Comparing SFS-AI with FRAX with BMD and BMD

#### Women of any age

Comparisons of the diagnostics was confined to participants having all three measurements. **Figure 1** shows the diagnostics as continuous traits. For ‘All Fragility Fractures’ and ‘Major Fragility Fractures’, the AUCs for SFS-AI were 72% and 69% respectively (p < 0.05). **Table S3** shows unadjusted and adjusted SFS-AI predicted women having either category of fractures (ORs ranged from 2.07 to 2.41, all p < 0.001). Neither of the other two diagnostics predicted either category of fracture. **Figure 1** also shows the diagnostics as categorical traits. For SFS-AI, sensitivities were 59.3% and 50.0% for detecting women having ‘All Fragility Fractures’ or ‘Major Fragility Fractures’ respectively, values that were significantly greater than sensitivities of FRAX with BMD or BMD (which ranged 4.2 to 16.7%). Specificities of SFS-AI were 77.1%, significantly lower than specificities of the other two diagnostics (which ranged 94.6 to 96.6%).

#### Women aged ≥ 65 years

Supplementary **Figure S3** shows the performance of the diagnostics as continuous traits. For ‘All Fragility Fractures’ and ‘Major Fragility Fractures’, the AUCs for SFS-AI were 70% and 66% respectively. **Table S4** shows unadjusted and adjusted SFS-AI predicted both categories of fractures (OR 1.68 to 2.15, all p < 0.05).

Neither of the other two diagnostics predicted either category of fracture. **Figure S3** also shows the performance of the diagnostics as categorical traits. For SFS-AI, sensitivities were 67.6% and 60.0% for detecting women having ‘All Fragility Fractures’ or ‘Major Fragility Fractures’ respectively, values significantly greater than the sensitivities of FRAX with BMD or BMD (ranging 6.67 to 26.7%). Specificities of SFS-AI were 70.7%, significantly lower than specificity of 94.6% for the other two diagnostics.

#### The morphological basis of the SFS-AI

**Figure 2** shows that SFS-AI increased across age in women having fragility fractures and in women remaining fracture-free. Red regions of the heat map overlying bone and soft tissue identify regions of high relevance to risk of incident fractures compared to the blue regions. **Figure 3** shows that SFS-AI correlated with microarchitecture; directly with cortical porosity and FRAX with BMD, and negatively with trabecular density and BMD. **Figure 3** and Supplementary **Table S5** show that 46% of the variance in SFS-AI was explained by variance in age (p = 0.002), cortical porosity and trabecular density (both p < 0.001) but not with BMD or FRAX with BMD; 54% of the variance remained unexplained.

## Discussion

A deep learning algorithm was trained to identify women having fragility fractures using only the high-resolution three-dimensional images of bone and soft tissue. No other information was used. When training no longer improved predictive strength, the algorithm was tested in a cohort without knowledge of their fracture status during the ensuing 5 years. This algorithm served as a surrogate of fracture risk, predicting the incidence of ‘All Fragility Fractures’ and ‘Major Fragility Fractures’ and did so in women 65 years and older with a sensitivity and specificity of 60-70%, out-performing BMD and FRAX with BMD, neither of which predicted fractures.

This surrogate of fracture risk, a Structural Fragility Score derived by deep learning artificial intelligence, increased across advancing age, was higher in women having incident fractures than those remaining fracture-free, and correlated directly with cortical porosity and negatively with trabecular density. Deterioration of these two traits produces a nonlinear increase in bone fragility,^22^ predicts incident fractures,^8,9^ prevalent fractures^40^ and predicts estimated bone strength independent of BMD.^41^ Deterioration of these two traits accounted for most of the 48% explained variance in SFS-AI. BMD was not an independent predictor of SFS-AI.

Many qualities of bone not captured by BMD but not yet quantifiable non-invasively, may contribute to the 54% of the unexplained variance in this surrogate of fracture risk.^14-19^ For example, heterogeneity in bone’s material composition forms discontinuities, edges, that defend against fracture by increasing the energy required to initiate and propagate a crack.^42^ Small changes in the degree of mineralization increase matrix stiffness but reduce its ductility (ability to absorb energy by deforming).^43^ Heterogeneity in the size and number of osteons,^44^ the cement line around each osteon,^45,46^ the differing orientation of mineralized collagen fibres of adjacent concentric osteonal lamellae,^47-49^ the extent glycation,^50^ hydration^51^ and other factors^52,53^ influence the mechanical properties of bone. The heat map implicated deterioration of soft tissue as well as bone. The nature of soft tissue deterioration is not known but if it is sarcopenia then the SFS-AI algorithm might capture a component of risk for falls.^23^

Most studies using machine learning are cross sectional and examine the ability to identify persons with prevalent fractures or osteoporosis (BMD T-Score ≤ - 2.5 SD).^24-27^ This is the first prospective study using deep learning to derive an algorithm that identifies women having incident fractures during five years. The algorithm was developed by interrogating the three-dimensional images of bone and soft tissue, no other information was used. This Structural Fragility Score serves as a surrogate of fracture risk that is likely to assist in reducing the population burden of fragility fractures. It provides a diagnostic able to identify most women at risk of fracture and provides fast processing, easy access to risk assessment allowing prompt initiation and monitoring of preventative treatment at the community level.

High resolution peripheral quantitative computed tomography (HRpQCT) technology is no-longer confined to the research domain. Commercial devices are now CE marked and FDA cleared for multiple clinical settings. Analysis requires only the acquisition of the three-dimensional image of the distal radius, ulna and soft tissue and cloud-based computer technology provides prompt diagnosis allowing initiation or monitoring of therapy.

This study has several limitations. Further studies are needed to determine whether including factors predisposing to falls such as muscle mass and function, age, height, weight and other covariates improves the performance of the diagnostic. The sample sizes were insufficient to evaluate performance of the diagnostic in predicting individual types of fracture.

Advancing age is accompanied by deterioration in bone mass, its material composition, architecture and muscle mass - factors contributing to fragility fractures, a public health problem. High-resolution quantitative computed tomography and deep learning provide a Structural Fragility Score that serves as a holistic surrogate of fracture risk. This diagnostic is an accurate, safe, rapid and easily accessible tool that captures the deterioration of bone qualities contributing to bone fragility independent of BMD and perhaps deterioration of muscle predisposing falls. This surrogate identifies women at high risk of fracture needing prompt treatment to avert fracture and may allow monitoring the success or failure of treatment.

## Data Availability

Data may be made available upon reasonable request for further analysis, after examination of an analysis proposal by the investigators.

## Legends for Figures in supplementary material

**Figure S1**. Structure of the deep learning model used to predict fracture. The input is the 110 slices of a wrist scan used to acquire the three-dimensional image of the distal radius, distal ulna and adjacent soft tissue. DenseNet121 is used as the neural network backbone. The output feature after the global average pool is a 256-dimension feature. A multi-task (age prediction, fracture prediction and non-fracture years prediction) learning strategy was used to achieve robust extraction of relevant features.

**Figure S2**. We studied women from OFELY (n = 568), Qualyor (n = 1427) and Gerico (n = 671). (A) There were 526, 1187 and 387 images remaining from the respective cohorts for analysis after excluding images from women remaining fracture-free followed for under 5 years and images from women having traumatic (non-fragility). (B) Women having a fragility fracture during 5 years were denoted as (+), women remaining fracture-free as (-). (C) Participants from each cohort were randomly allotted into five groups with approximately equal numbers of (+) and (-) subjects. See Methods section.

**Figure S3. Left two panels**. Receiver Operator Characteristic (ROC) curves for Structural Fragility Score Artificial Intelligence (SFS-AI), Fracture Risk Assessment Score (FRAX) with bone mineral density (BMD) and BMD as continuous traits predicting ‘All Fragility Fractures’ and ‘Major Fragility Fractures’ in women 65 years of age and over. Area under the Curves (AUCs) with 95% Confidence Intervals (CI) were significantly different from 0.5 (*p < 0.05) for SFS-AI only. **Right two panels**. Sensitivity and specificity of the SFS-AI, FRAX with BMD and BMD as categorical traits predicting women having ‘All Fragility Fractures’ or ‘Major Fragility Fractures’.

## Supplementary AI methods

### Deep Learning Network, training and the heat map

DenseNet used dense connection between layers and achieved efficacy in feature extraction. The first layer had a 110-dimension input instead of 3-dimension input in the original DenseNet.^34^ We added a transition layer using 1*1 convolution after the last denseblock of DenseNet to reduce the feature dimension from 1024 to 256 to extract a compact feature. A multi-task learning strategy was used to achieve more robust features. Extracted features were used to predict fractures during the ensuing 5-years, to predict the patient’s age at the time of scanning and the duration of the fracture-free years of follow-up since scanning. The latter two tasks are included in the training to produce generalized feature representations powerful enough to be shared across different tasks.

We used a pretrained model as the initial model and 0.5 as the classification threshold for the training and validation. During training, cross-entropy loss was used as the fracture situation prediction loss, and used the mean squared error loss as the loss of age prediction and non-fracture year prediction. L1 regularization is performed on the weights of the classification layer to reduce overfitting. The model is optimized by ADAM optimizer using the four losses with weight 1 on the first three losses and weight 0.01 on the L1 regularization loss.^35^ The learning rate is set as 5e-6 for the DenseNet backbone and 5e-5 for the other layers, including the transition layer and the three multi-learning branches. Pytorch is chosen to implement the model training and testing.^36^ The Grad-CAM is utilized to generate the heatmap to represent the features that are extracted by the deep learning model.^37^

